# Parent-reported phenotype data on chromosome 6 aberrations collected via an online questionnaire: data consistency and data availability

**DOI:** 10.1101/2022.11.07.22282039

**Authors:** Aafke Engwerda, Barbara Frentz, Eleana Rraku, Nadia F. Simoes de Souza, Morris A. Swertz, Mirjam Plantinga, Wilhelmina S. Kerstjens-Frederikse, Adelita V. Ranchor, Conny M. A. van Ravenswaaij-Arts

## Abstract

**Background:** Even with the introduction of new genetic techniques that enable accurate genomic characterization, knowledge about the phenotypic spectrum of rare chromosomal disorders is still limited, both in literature and existing databases. Yet this clinical information is of utmost importance for health professionals and the parents of children with rare diseases. Since existing databases are often hampered by the limited time and willingness of health professionals to input new data, we collected phenotype data directly from parents of children with a chromosome 6 disorder. These parents were reached via social media, and the information was collected via the online Chromosome 6 Questionnaire, which includes 115 main questions on congenital abnormalities, medical problems, behaviour, growth and development. Here, we assess data consistency by comparing parent-reported phenotypes to phenotypes based on copies of medical files for the same individual and data availability by comparing the data available on specific characteristics reported by parents to data available in existing literature.

**Results:** The reported answers to the main questions on phenotype characteristics were 85–95% consistent, and the consistency of answers to subsequent more detailed questions was 77–96%. For all but two main questions, significantly more data was collected from parents via the Chromosome 6 Questionnaire than was currently available in literature. For the topics developmental delay and brain abnormalities, no significant difference in the amount of available data was found. The only feature for which significantly more data was available in literature was a sub-question on the type of brain abnormality present.

**Conclusions:** This is the first study to compare phenotype data collected directly from parents to data extracted from medical files on the same individuals. We found that the data was highly consistent, and phenotype data collected via the online Chromosome 6 Questionnaire resulted in more available information on most clinical characteristics when compared to phenotypes reported in literature reports thus far. We encourage active patient participation in rare disease research and have shown that parent-reported phenotypes are very reliable and contribute to our knowledge of the phenotypic spectrum of rare chromosomal disorders.

## Background

Knowledge about rare diseases is often limited. This leads to a lack of critical information, including information on prognosis, co-morbidity, treatment options and surveillance, that is essential to health professionals and parents of children with these rare disorders. Facing this gap, parents of children with rare disorders often start searching the internet, and many now end up uniting via social media platforms in their quest to find more information on the effects and prognosis of their child’s disorder.

Recently, Miller et al (2021) reviewed the use of social media in research on rare genetic diseases. They showed that there has been a rapid increase in social media use for rare disease research over the past 13 years. Social media was mainly used for recruitment of study participants and primary data collection (including surveys hosted on a different platform), and most researchers used Facebook or Twitter. They concluded that social media can be a powerful tool in studying rare disease populations^1^.

Parents of children with a rare chromosome 6 aberration also came together in an international Facebook support group. Since they did not have the scientific knowledge and expertise to study rare chromosomal disorders, they approached the Center of Expertise for Rare Chromosome Disorders at the University Medical Center Groningen (coordinated by author CR) with a request to study the effect of the aberrations found in their children. This was the start of a collaboration that resulted in the Chromosome 6 Project, which is “Driven by parents, for parents”. Over time, the closed Facebook group grew to over 1200 members, and more than 250 families worldwide currently participate in the phenotype–genotype studies of the Chromosome 6 Project^2–5^. The aim of this project is to collect detailed and accessible information on the effect of chromosome 6 aberrations and to provide this crucial information to parents and health professionals.

Most chromosome 6 aberrations are unique, with only a few individuals worldwide sharing a comparable aberration. With the introduction of the very precise and reproducible microarray technique in routine diagnostics in 2004^6^, it became possible to compare and study rare chromosomal aberrations in a highly reliable way, worldwide. Nonetheless, the available information on individuals with rare chromosomal disorders remains very limited. Existing databases, such as DECIPHER (deciphergenomics.org) ^7^, rely on the time and willingness of health professionals to enter information. Moreover, information in published case reports is often limited and focused on a specific topic of interest to the authors, for example brain abnormalities. The level of detail on the addressed topics is often high in these papers, but the full phenotype is not always described. Yet a full phenotype description enhances the identification of new characteristics, for example those with a low penetrance, and helps to unravel the full phenotypic spectrum of a rare disorder.

The Chromosome 6 Project collects phenotype data directly from parents via the multilingual^2^ online Chromosome 6 Questionnaire^2^. Parents complete this questionnaire based on their own experiences and knowledge, including information they received during medical consultations. By collecting data directly from parents, most of whom are highly motivated to participate, we are not dependent on the limited time health professionals can devote to sharing this kind of data. Since the number of individuals using social media keeps increasing^8^, unlocking this source of parent-reported data will help us gain insight into the phenotypic spectrum of rare chromosomal disorders. However, questions may arise about the quality of data provided by parents.

In this study we sought to answer two questions. First, is the phenotype data provided by parents consistent with what can be extracted from their child’s medical files? Second, are there differences in the availability of data on specific phenotypic features presented in literature case reports as compared to data collected from parents through the Chromosome 6 Questionnaire? The current study explores the usability and added value of data collected directly from parents of children with a rare chromosomal disorder in order to enhance our knowledge on its phenotypic spectrum. This is important for future studies on rare diseases and encourages patient participation.

## Methods

To examine data consistency, we compared phenotype data collected from parents for 20 children recruited through the Chromosome 6 Project to data from their child’s medical files. To explore data availability, we compared data on 34 individuals from the Chromosome 6 Project with data on 39 individuals with comparable aberrations reported in literature.

Recruitment and data collection within the Chromosome 6 Project was previously described in detail by Engwerda et al. 2018^2^. In short, information about the Chromosome 6 Project was shared via Facebook (Chromosome 6 Facebook group), Twitter (@C6study) and the project’s website (www.chromosome6.org). Patients or their legal representatives, from here on called “parents”, could sign-up via our website in order to consent to and participate in the research. Inclusion criteria were an isolated chromosome 6 aberration and a microarray report, which was requested during sign-up. After sign-up, parents received a personal account to access the multilingual online Chromosome 6 Questionnaire that was used to collect the phenotype data based on the MOLGENIS platform^9^. The questionnaire consists of questions about the pregnancy and birth, congenital abnormalities, relevant dysmorphic features, development (including developmental milestones derived from the ‘Van Wiechenschema’, a developmental scale for children used by most Dutch health professionals^10^, and questions on adult functioning derived from the Lawton Instrumental Activities of Daily Living (IADL) Scale^11^), behaviour and health-related problems of an individual. The questionnaire is tailored to previously given answers, with only applicable questions shown based on age, sex and other previously given answers. Parents used different sources of information to complete the questionnaire: personal observations and/or experience, oral or written information from health professionals and the (electronic) patient file if they had access to it. For less than 10% of the completed questionnaires, the adult individual with the chromosome 6 aberration rather than their parents completed the questionnaire. These individuals also included parents who had the same aberration as their child.

To answer our two research questions, we focused on the answers to 115 of the 132 main questions in the Chromosome 6 Questionnaire. These questions were selected based on their clinical relevance for describing the phenotype of individuals with chromosomal aberrations. For example, questions on the presence of seizures and a formal diagnosis of epilepsy were included, but questions on the number of seizures were excluded. In addition, we looked at 71 sub-questions that were only asked based on previously given answers. Almost all the main questions in the Chromosome 6 Questionnaire are mandatory. The only exception is questions on growth measurements, as not all parents keep a history of growth measurements for their child and we ask for measurements at different points in time (if applicable): birth, current age and the ages of 1, 2, 4, 6, 9, 12 and 16 years. To study whether there were differences between different topics covered by the questionnaire, questions were grouped in five categories: congenital abnormalities, medical problems, behaviour, growth and development. Within these five categories, the questions were further divided into subcategories. See Supplementary Information S1, Additional File 1, for the included questions and the category and subcategory definitions.

### Data consistency: parent-reported data versus medical file data

We performed a data consistency study to analyse whether phenotype data collected directly from parents (parent data) is comparable to data collected from treating physicians (medical file data).

Forty-three parents who had previously participated in a pilot study were invited via email to test the newly developed online Chromosome 6 Questionnaire in English. The phenotype data collected through these parents was planned to be used as the ‘parent data’ for the data consistency study. Selection criteria for being on the test panel were: being a parent of a child with a chromosome 6 aberration, living in an English-speaking country and the availability of copies of medical files received from the treating physicians. The parents were informed that the data submitted to the questionnaire would not be destroyed after the test phase and would be used for future genotype–phenotype studies on the chromosomal aberration of their child. The parents received a personal account for the online Chromosome 6 Questionnaire via email and were asked to complete the new questionnaire as precisely as possible and provide comments for improvements.

Of the 43 parents invited, 20 did not respond to the invitation or reminder email and another 3 parents informed us that they could not be part of the test panel due to time constraints. Twenty parents (47%) completed the questionnaire for their child, and these completed questionnaires are the source for the ‘parent data’ in our data consistency study (see Figure S1, Additional File 1). The parents in the test panel came from six countries: Australia, Canada, Ireland, New Zealand, the United Kingdom and the United States of America, see Table S1, Additional File 1. Further details on the characteristics of the 20 parents and their children can be found in Table 1.

**Table 1.**
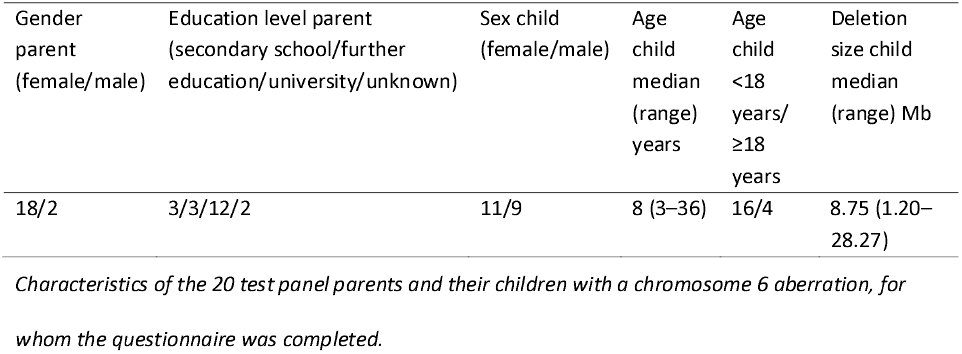
Characteristics of the 20 test panel parents and their children

### Medical file data

As mentioned above, a pilot study was performed before the introduction of the online Chromosome 6 Questionnaire. As part of this pilot study, informed consent was obtained to request copies of the medical files from the treating physicians of participants with a chromosome 6 aberration. For the 20 children of the parents in our test panel, copies of parts of the medical files had been received from up to 11 different treating physicians. For most individuals, we had data from a geneticist, neurologist and/or paediatrician. Details on the medical files are presented in Supplementary Tables S2 and S3, Additional File 1. The copies of the medical files were the source for the ‘medical file data’ in our data consistency study. A physician in the project group, who was not involved in the child’s care, completed a duplicate of the Chromosome 6 Questionnaire for each individual using only the copies of the medical files. This resulted in data in the same format, enabling comparison of the ‘parent data’ and ‘medical file data’. If multiple medical files were available, the most recent information was used to complete the duplicate of the questionnaire.

### Data analysis

The answers from the parent data and medical file data obtained via the Chromosome 6 Questionnaire were compared for consistency for each individual. The data was defined as consistent when the answers were the same and as *inconsistent* if there was a discrepancy between the answers. However, not all data could be directly compared in this way, and some alterations were made to enable comparison. Growth measurement data was converted to standard deviations (SDs) and was defined as consistent if there was a ≤1 SD difference between the two data sources and inconsistent if there was a >1 SD difference between the sources at a specific age. For some individuals, growth measurements were available for multiple time points from both sources (parent data and medical file data). If this was the case, the youngest and oldest ages for which a measurement was available were checked for consistency. We also counted the number of available measurements for both sources (Table S4, Additional File 1).

For some sub-questions, multiple answer options could be chosen (e.g. different types of heart defects). These multiple-answer options were considered as one answer for this question and defined as partly (in)consistent if one part of the given answers was consistent and another inconsistent. Answers to sub-questions on developmental delay included IQ test scores and age of achievement for developmental milestones, with the latter converted to developmental quotients (DQ) as described previously^4^. The IQ test scores and DQs were categorized as normal (>85), borderline (70–85), mild (50–70), moderate (30–50) or severe (<30) delay. Answers were considered consistent when there was ≤1 category difference and inconsistent when there was >1 category difference.

### Data presentation

The degree of consistency for answers is presented as: consistent/observed x 100 = consistency, in which observed = consistent + inconsistent. Data could only be compared for consistency if it was available for both data sources. If the answer to a main question (n=115) was known for one data source but not the other, the answer could not be checked for consistency. The frequency of an answer being unknown was counted for both data sources (Table S5, Additional File 1). This showed that for 99.6% (1536/1542) of the compared answers, an answer was unknown from the medical file data but we did have an answer from the parent data. Taking into account that characteristics absent in an individual are often not reported in the medical files, we considered an answer “probably consistent” if a characteristic was reported as absent in the parent data and unknown (absent and unknown) in the medical file data. By doing this, we could also calculate a degree of probably consistent answers: (consistent + (absent and unknown)) / (observed + (absent and unknown)) x 100 = probably consistent.

Since the answers to sub-questions can be partly consistent, the degree of consistency and the degree of partial consistency are presented.

We categorized inconsistent answers as *major* inconsistent, *minor* inconsistent or *partly* inconsistent (as explained above). Inconsistent answers for main questions were considered major. Inconsistent answers for sub-questions were considered minor. Inconsistent answers to seven main questions (marked with an * in Supplementary Information S1, Additional File 1) were categorized differently because the answers to these questions depend on each other. For these questions, an inconsistent answer was considered minor.

We studied the degree of consistency and major and minor inconsistencies for the total number of answers, for specific categories and subcategories of questions (i.e. congenital abnormalities and brain abnormalities) and for the 20 parents in the test panel.

### Data availability: parent data cohort versus literature data cohort

Data collected on individuals with terminal 6q deletions was used for the data availability study. Individuals were selected from the Chromosome 6 Project and from literature using previously described criteria^2,4^ plus two additional criteria: foetuses and new-borns were excluded and only index cases (or the person best-described in the case report) were included. The Chromosome 6 Questionnaire was completed by parents for their child and by a researcher of the Chromosome 6 Project for the literature cases. Details on completing the Chromosome 6 Questionnaire for literature case reports were described before^2^. Data on individuals collected from parents formed the parent data cohort and data on individuals described in case reports formed the literature data cohort. Selection of individuals based on the inclusion criteria resulted in 34 individuals in the parent data cohort and 39 individuals in the literature data cohort (Figure S2, Additional File 1). Details on both cohorts are provided in Table 2.

**Table 2.** Characteristics of the parent data cohort and literature data cohort

|  | Sex<br>(female/male) | Age median (range)<br>years; months | Deletion size median<br>(range) Mb |
| --- | --- | --- | --- |
| Total (n=73) | 47/26 | 6;0 (0;4–37;0) | 7.53 (0.39–16.00) |
| Parent data cohort (n=34) | 26/8 | 4;6 (0;6–32;9) | 8.44 (0.76–16.00) |
| Literature data cohort (n=39) | 21/18 | 8;0 (0;4–37;0) | 5.79 (0.39–13.15) |

*Characteristics of individuals included in the parent data cohort and the literature data cohort. More details are provided in Engwerda et al 2022.<sup>4</sup>*

### Data analysis

Data was classified as *available* (feature present or absent) or *unavailable* (feature unknown or missing). For follow-up, growth measurement data was classified as available when at least one measurement was available. For main questions, significant differences in the availability of data for the two cohorts was tested using a Chi-square test and a Fisher’s exact test when applicable (more than 20% of the expected counts below 5). For sub-questions, a Fisher’s exact test was used because less data was available.

## Results

### Data consistency

The percent data consistency for the total number of answers and for the five topic categories is listed in Table 3. The data for the main questions, including all subcategories, is visualized in Figure 1. The absolute numbers and details on how often answers were unknown for the parent data and medical file data can be found in Supplementary Tables S5 and S6, Additional File 1.

**Table 3.**
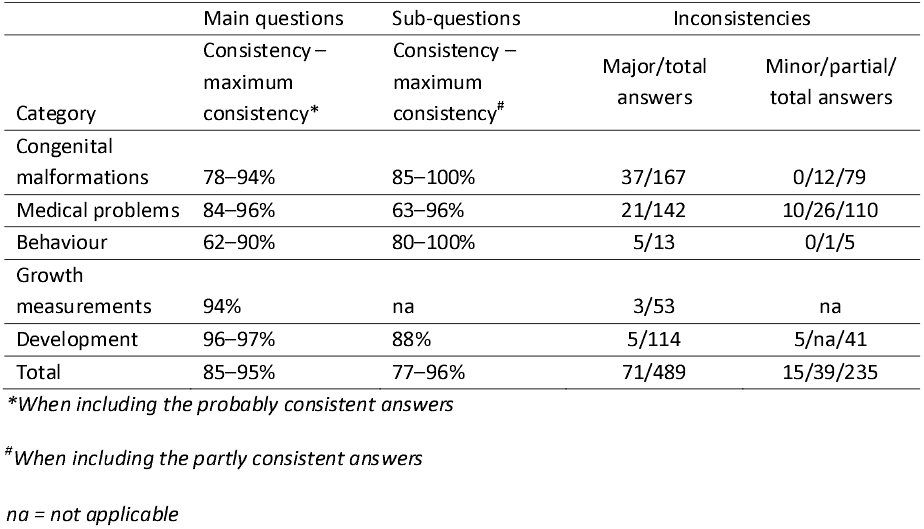
Summary data consistency and inconsistencies per question category

**Figure 1.**
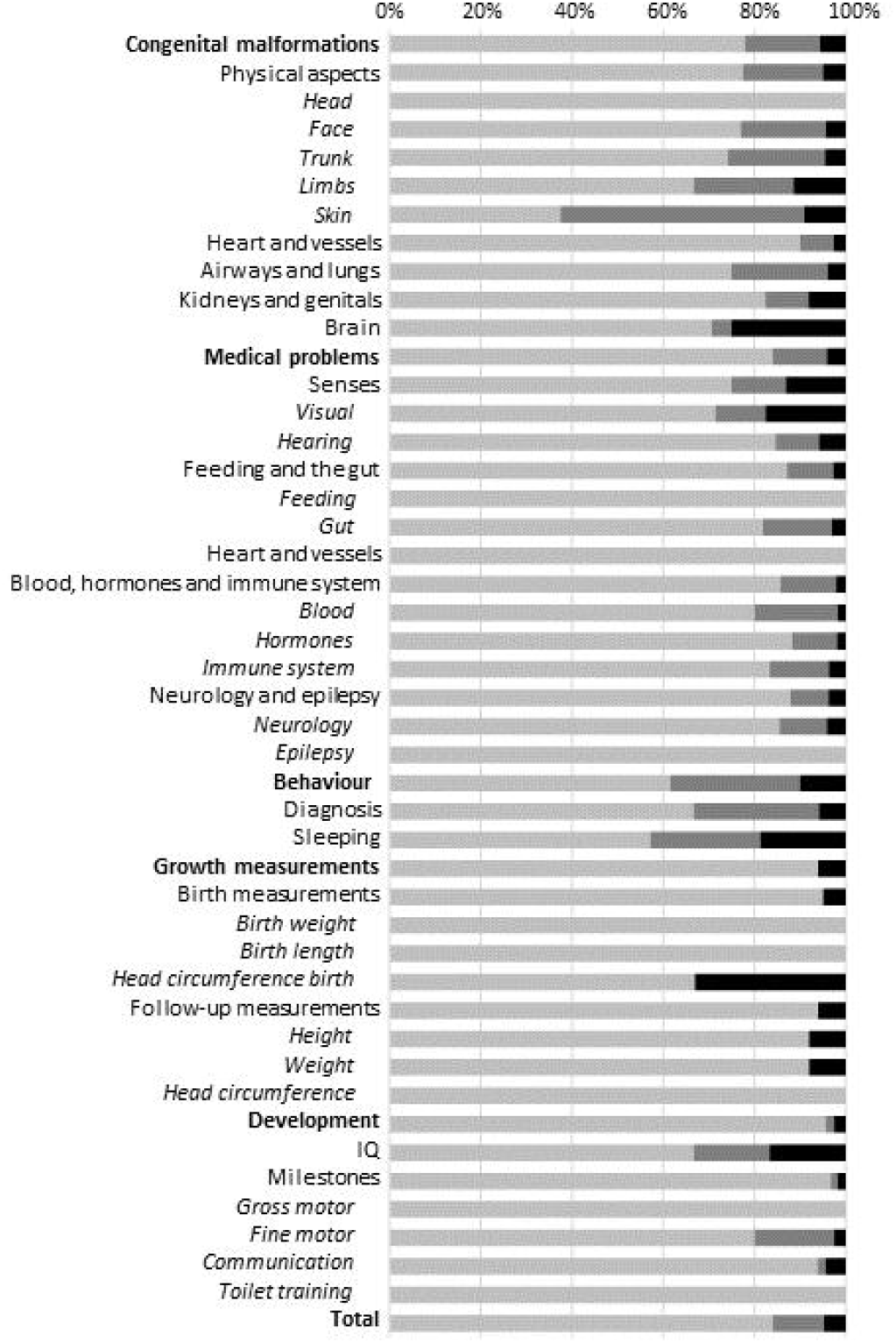
Data consistency for main questions. Light grey: percentage of consistent answers for each (sub) category. Darker grey: the maximum consistency when probably consistent answers were taken into consideration. Black: percentage of inconsistent answers.

### Details in Supplementary Table S5 and S6, Additional File 1, and visualized in Figure 1

For the main questions, the consistency of the collected answers could be compared for 25% (521/2063) of the answers, and consistency was 85% (Tables 3 and S5, Additional File 1). In total, 1542 answers (75%) could not be compared for consistency due to lack of data in one of the two data sources, predominantly the medical files (99.6%, 1536/1542) (Table S5, Additional File 1). By taking into account that unknown answers in the medical file data were probably absent characteristics, and considering these answers were probably consistent when parents reported the characteristic to be absent, we could check an extra 1119 answers for consistency (Table S7, Additional File 1). The percentage of answers that could be compared for consistency then increased from 25% to 79% (1640/2063) and resulted in a maximum consistency of 95% for all main questions.

There were differences in the level of consistency among the question categories (Figure 1). Consistency was highest for questions about development (96–97%) and lowest for those about behaviour (62–90%). Within the category of congenital malformations, answers to main questions on brain (71–75%, 12/17–15/20), limb (67–89%, 14/21–54/61) and skin (38–91%, 3/8–50/55) abnormalities were the least consistent and data on dysmorphic features of head shape (100%, 29/29–91/91) was the most consistent. In the category medical problems, answers were fully consistent for questions on feeding (100%, 9/9–13/13), heart and vessels (100%, 1/1–39/39) and epilepsy (100%, 11/11–18/18). Answers to questions in the subcategory visual, including vision and eye problems, were the least consistent (71–82%, 25/35–47/57).

For answers to main questions on development, consistency was high for the achievement of milestones (97–98%, 88/91–165/168) and fully consistent for gross motor milestones (100%, 52/52– 61/61) (Table S5, Additional File 1). Consistency was lowest for answers on questions regarding IQ (91–93%, 21/23–27/29). For two individuals, parents reported that developmental testing was not performed, while the contrary was reported in the medical files. Four individuals in our cohort reached adulthood (age >18 years) (Table 1). For three of them, the parent data contained answers for the eight questions on adult functioning. For two of the four individuals, copies of the medical files at age >18 years were available, but data on adult functioning was available for only one individual. For this individual, two answers could be checked and were consistent (data not shown).

For the sub-questions, consistency was 77% (Tables 3 and S6, Additional File 1). If partly (in)consistent answers were considered consistent, overall consistency rose to 96%. For some subcategories with multiple answer options, such as brain abnormalities, consistency increased substantially when partially consistent answers were included in the consistency range (see Discussion). Consistency for answers to sub-questions was highest for congenital malformations (85– 100%) and lowest for development (88%).

In total, 71 major and 15 minor inconsistencies were identified (Table S8, Additional File 1). Most major inconsistencies (52/71, 73%, data not shown) were for answers on characteristics reported to be present in the medical files but reported absent by parents. For example, for the subcategories face, limb and skin abnormalities, an abnormality was reported in the medical files 6, 7 and 5 times, respectively, whereas the parents did not report an abnormality. When the subsequent answers to the sub-questions were studied, these abnormalities included hypertelorism, a cleft soft palate, a lip pit, generalized joint hypermobility, pes planus, capillary skin haemangiomas and café au lait spots. In 23% (16/71, data not shown) of the major inconsistencies, parents reported a characteristic to be present that was absent based on the medical files. Abnormalities reported by parents included, amongst others, a congenital heart defect (atrial septal defect), renal or urinary tract abnormality (not defined), abnormal curvature of the spine (scoliosis), anaemia, mild and moderate hearing impairment and sleeping problems (insomnia). Lastly, 3/71 (4%, data not shown) major inconsistencies were found in the growth measurement data.

Two subcategories stand out for the number of major inconsistencies: brain and vision. The highest number of inconsistent answers, five answers, was seen in the subcategory brain. In two cases, the parents reported a brain abnormality (an intracranial cystic lesion, hydrocephalus and corpus callosum abnormality in one individual and undefined brain abnormalities in the other), while brain abnormalities were reported to be absent in the medical files. In the other three cases, the medical files reported a brain abnormality (cortical dysplasia and ventriculomegaly in one individual, hydrocephalus in another and brain abnormalities not further defined in the third), while the parents did not. For the category vision, ten answers were inconsistent. For two answers, a characteristic was reported as present by the parents (cortical visual impairment and mildly reduced visual acuity) but not reported in the medical file, whereas the doctor reported a characteristic to be present (including mildly reduced visual acuity, strabismus and oculomotor apraxia) but the parent did not for eight answers.

Five minor inconsistencies were the result of an abnormality reported to be present in the medical file data and absent in the parent data (formal diagnosis of epilepsy, hypotonia, areflexia), and five were present in the parent data but absent in the medical file data (including status epilepticus, hypotonia, hyperreflexia and areflexia). The last five minor inconsistencies were found in sub-questions on the ages of achievement for milestones, predominantly gross motor milestones.

Lastly, we also examined the overall consistency of the answers per parent in the test panel. The median (range) number of answers that could be checked for consistency was 36 (17–59) per parent. The consistency was 83% (71–100%), and the median number of inconsistent answers per parent was 5.5 (0–15). Subdivided for major, minor and partial inconsistencies, this was 3 (0–8), 0 (0– 3) and 1.5 (0–7), respectively. See Table S9, Additional File 1, for the distribution per parent and Figures 2 and S3, Additional File 1, for a visualization. The parent for whom most answers could be checked for consistency also gave the most inconsistent answers in absolute numbers.

**Figure 2.**
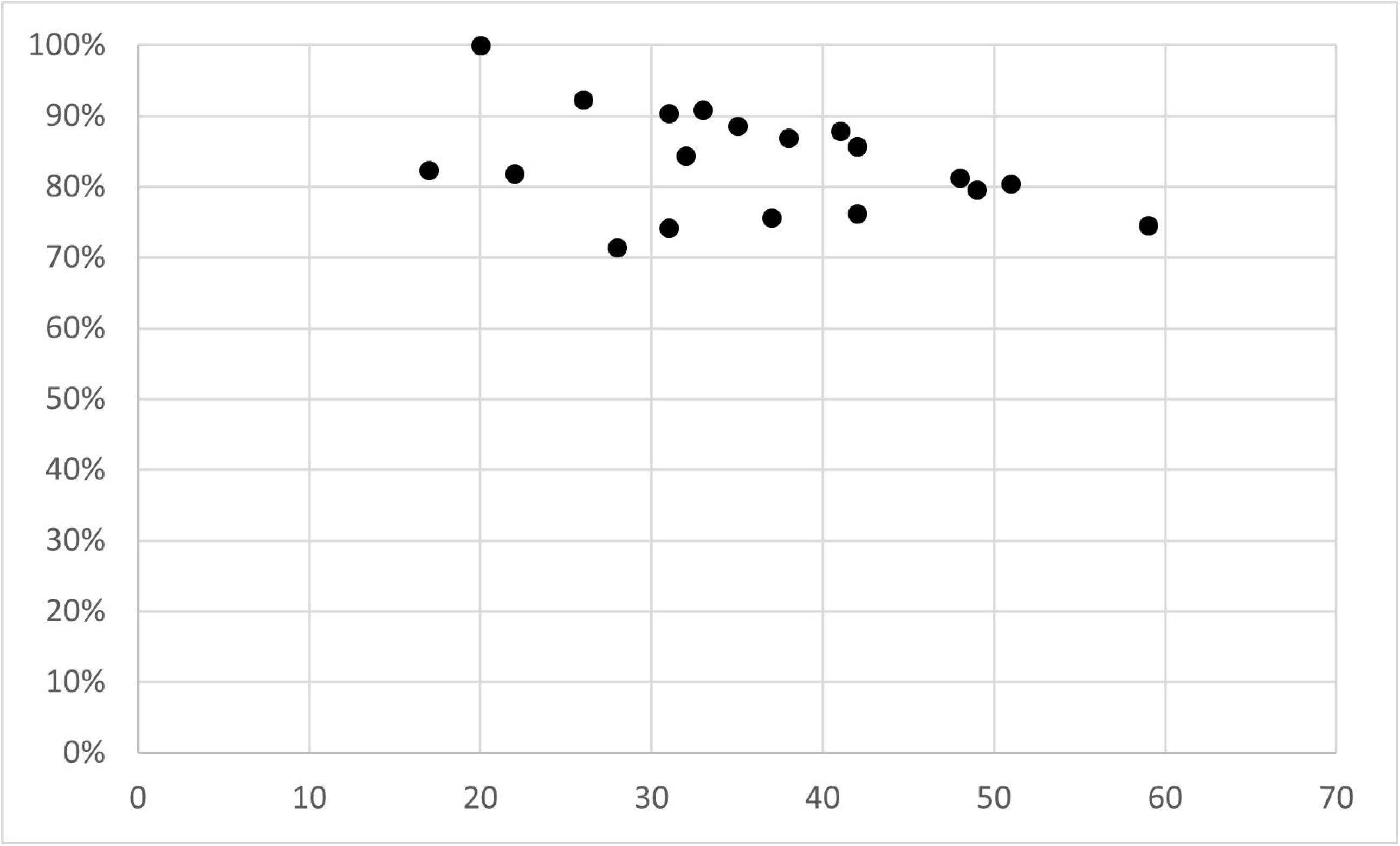
Data consistency for 20 test panel parents. Each dot represents one parent in the test panel (n=20). X-axis is the number of answers that could be checked for consistency. Y-axis is the percentage of consistent answers. For details and absolute numbers, see Table S9, Additional File 1.

### Data availability

Table 4 summarizes for how many questions a significant difference was found in the availability of data collected from parents compared to literature case reports. Notable p-values are presented in Table 5. See Tables S10–S12, Additional File 1, for details on significance per subcategory and the p-values for all questions.

**Table 4.** Summary of the availability of data collected from parents as compared to literature

|  | Main questions | Sub-questions |
| --- | --- | --- |
| | Significantly more data from parents/significantly more data from literature/no significant difference ($\alpha=0.05$) | Significantly more data from parents/significantly more data from literature/no significant difference ($\alpha=0.05$) |
| Congenital malformations | 37/0/1 | 1/1/24 |
| Medical problems | 38/0/0 | 2/0/19 |
| Behaviour | 3/0/0 | 0/0/3 |
| Growth measurements | 6/0/0 | na |
| Development | 29/0/1 | 2/0/19 |
| Total | 113/0/2 | 5/1/65 |

**Table 5.**
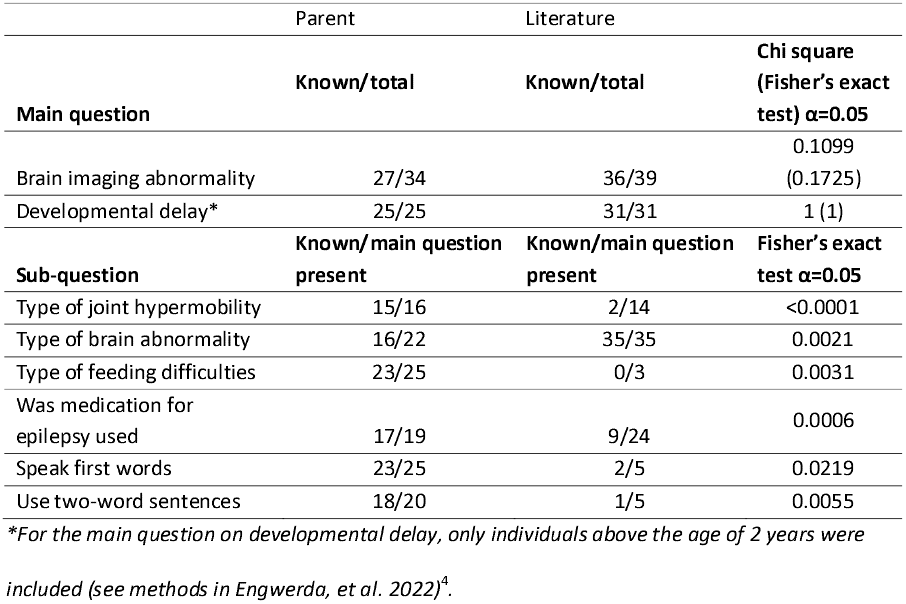
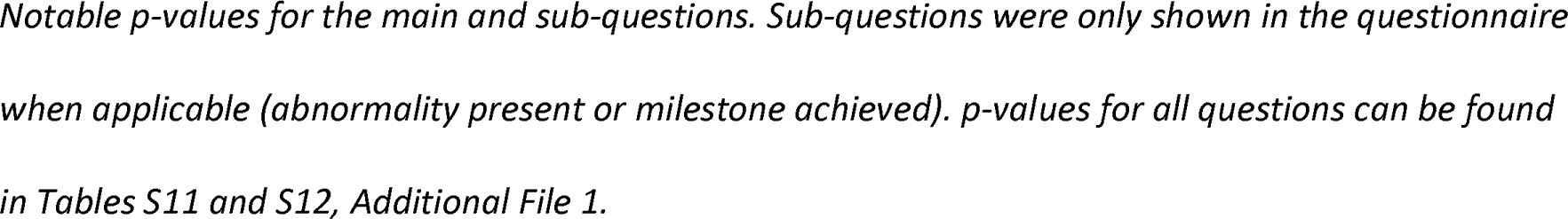
Notable p-values of data availability for main and sub-questions

For 113 of the 115 main questions, a significant difference in data availability was found in favour of the parent data. The two questions for which no significant difference was found were on the presence of brain abnormalities (Fisher’s exact test, p=0.1725) and the presence of developmental delay (Fisher’s exact test, p=1) (Table 5). For 65 sub-questions, no significant difference was found in the availability of data on the type of abnormality or, for milestones, the age of achievement. For six sub-questions, a statistically significant difference was found, as listed in Table 5. Significantly more information was available from parents about the type of joints affected in hypermobility, the type of feeding difficulty, medication use for epilepsy and the age of achievement of the milestones “speak first words” and “use two-word sentences”. In contrast, more information was available from literature about the type of brain abnormalities.

## Discussion

### Data consistency

To the best of our knowledge, this is the first study to compare parent-reported phenotype data to data derived from medical files on the same individual. Most answers reported by parents were consistent with the answers collected from the medical files. We found 85–95% consistency for the main questions and 77–96% consistency for sub-questions (Table 3).

While we observed major and minor inconsistent answers in all categories of questions, no inconsistencies were identified for most questions, and when inconsistent answers were identified, there were often only one or two per question. This suggests that no individual question was more susceptible to inconsistencies than another. The only exception to this was the question on whether brain abnormalities were present on imaging, which had five inconsistent answers.

The inconsistencies found could be explained by a number of factors. First, there was a difference in timing as the parent data was collected 2 years after the medical files were requested. New diagnoses, new medical problems and new milestones may have occurred in these 2 years that were reported by parents but lacking in the medical files. Second, characteristics reported as present by health professionals but absent by parents can reflect health professionals documenting all characteristics present but only informing parents about the clinically relevant ones. In this scenario, parents are not familiar with these characteristic because they were never communicated. We instructed the parents to fill out ‘absent’ if they had not heard a term before, since in most cases their child would probably not have that characteristic. Third, clinically less relevant characteristics could have been forgotten by parents as they had little effect on the clinical management, especially if other characteristics were more clinically pressing. To improve knowledge on prognosis, co-morbidity, treatment options and surveillance for rare disease patients, it is most important to collect information on clinically relevant characteristics.

Due to missing data, not all answers could be checked for consistency. To increase the number of answers to the main questions that could be checked, we considered the answer combination of “unknown characteristic” in the medical file data and “absent characteristic” according to the parent data to be “probably” consistent, as characteristics that are absent are often not reported in medical files. In contrast, for 390/1509 (26%) of the characteristics that were unknown according to the medical files, parents reported a characteristic to be present, possibly indicating that the information in the medical files we received was incomplete. Most of these missing answers (61%, 239/390) were in the category development and most concerned questions on achievement of milestones (see Supplemental information S1 and Table S8, Additional File 1). Data on achievement of milestones is often lacking from medical files if a detailed developmental report is not included, which explains the data missing from the medical files on this specific topic.

For sub-questions, we also took partly consistent answers into consideration. These were often seen for the sub-questions on brain abnormalities and feeding problems. This might be related to the fact that a larger number of answer options was available for these two questions (14 and 9, respectively). If a part of the answer was not the same for both data sources, the total answer was defined as partly consistent rather than completely inconsistent. We did this because one can debate whether it is clinically relevant if not all subtypes of brain abnormalities present in a child are reported by parents in a group of patients with extremely rare disorders for whom little is known on their overall phenotype.

If we had collected data solely from parents, the most clinically relevant information we would have missed were: vision and eye problems in eight individuals, brain abnormalities in three individuals and two formal diagnoses of epilepsy. On the other hand, parents also reported clinically relevant information that was not reported in the medical files, including one atrial septal defect, two individuals with hearing impairment, two with vision and eye problems and two with brain abnormalities. Since we do not know whether the medical file data were better than the data collected from parents, or the other way round, we can only conclude that in both cases we would have missed some clinically relevant data. This missed data may change the prevalence of certain characteristics. Nonetheless, our goal is not to prove that a clinical characteristic is seen in a certain percentage of individuals but to gain more knowledge and capture the spectrum of the full phenotype.

### Data availability

In our data availability study, we demonstrate that the full phenotype is not presented in most literature reports. For most main questions of the Chromosome 6 Questionnaire, the data collected from parents was more complete than the data extracted from literature case descriptions. This difference was expected, as parents were required to answer all the main questions on the questionnaire, meaning that data from parents was almost always available. Data collection from parents also resulted in relevant extra information, for example on sleeping problems. In the parent cohort, data on sleeping problems was available for 33 out of 34 individuals but was available for only 2 out of 39 individuals in the literature cohort. Notably, 50% of individuals with a terminal 6q deletion were reported to suffer from sleeping problems, which can be a major burden to parents ^4^.

No significant difference was found for two main questions: whether brain abnormalities were present (Fisher’s exact test, p=0.1725) and the question on the presence of developmental delay (Fisher’s exact test, p=1). This was also not surprising as the clinical characteristics most often reported in individuals with a terminal 6q deletion are brain abnormalities (reported in 91%) and developmental delay (reported in 92%) ^4^.

In the questionnaire, we collected data on the specific type of abnormality via sub-questions and saw little difference in data availability for these questions between parent and literature data. For five questions, more data was available from the parent data, but for the type of brain abnormalities this information was always available from the literature but only available for 73% of individuals in the parent data cohort. This might be explained by the high prevalence of brain abnormalities in terminal 6q deletions, leading to greater attention being paid to this abnormality in the literature ^4^. In the end, as discussed above, our goal is to collect data on the full phenotypic spectrum and not on exact prevalence numbers for a specific clinical characteristic. Overall, collecting data from parents led to more information on most characteristics when compared to literature, with parents only providing equivalent or less information for a few characteristics.

Recently, Li et al. reported on their online self-phenotyping tool, which resulted in the identification of underrepresented characteristics in literature and newly reported characteristics for 16p13.11 microduplication syndrome. These characteristics included, amongst others, aggression, anxiety, sleep disorders, feeding difficulties and immune disorders. Nineteen participants used the self-phenotyping tool to search and select characteristics applicable to their child or themselves ^12^. Using this tool, only characteristics that were present were registered. Characteristics known to be absent were not registered. Our Chromosome 6 Questionnaire contains 132 mandatory main questions, which results in the registration of characteristics being present and absent. Alongside the 115 main questions included here, the Chromosome 6 Questionnaire also contains main questions on more parent-experienced topics. For example, next to a question on formal behavioural diagnosis, which was included in the analysis in this paper, there is also a question on characteristics best describing their child’s behaviour. This question includes the following answer options (amongst others): being social, quiet, easily upset, showing self-harming behaviour and hyperactivity. At the end of the Chromosome 6 Questionnaire is an open text field in which characteristics not yet covered can be reported, giving parents the opportunity to report new characteristics.

### Limitations

Collecting phenotype data from parents via the online Chromosome 6 Questionnaire produces good quality data that is highly consistent with data from medical files and leads to more information on the full phenotypic spectrum than is available from literature reports. Nonetheless, there are also some limitations to our study. Firstly, we used social media to reach study participants for the Chromosome 6 Project. Although social media can reach people worldwide, it will only reach those with access to the internet and the knowledge and skills to participate in online social networking sites ^13^. Secondly, even though chromosome 6 aberrations often arise de novo and are not expected to be seen more often in certain populations, the availability of detailed genetic testing is not evenly distributed around the world. This means that individuals who cannot get a microarray report because of lack of access to genetic services cannot participate in the Chromosome 6 Project. Thirdly, only individuals from English-speaking countries were included in the data consistency study, which may have led to an ascertainment bias. For this study we were able to include 20 parents out of the 43 invited. However, these 43 parents, who had participated in the pilot study, were the early adopters and probably already the most active people in the social media community, while the selected 20 further had time and were willing to help develop and improve the questionnaire. This might have resulted in an overestimation of the consistency of the data provided by these active parents. For these reasons, we encourage future studies on data consistency that compare parent-reported data to health professional-reported data for the same individual, especially in the context of different rare disease populations.

### Implications

Despite the limitations of our study, phenotype data reported by parents will have a positive impact on both our knowledge of the phenotypic spectrum of a disorder and on future research. When comparing parent data and medical file data, some information was missing in both data cohorts. This suggests that combining parent data with medical file data would create the most complete dataset. However, the success of this endeavour will depend on the time and willingness of health professionals, and it will overshoot the goals we have set. In the end, this two-way approach will probably only further delay the availability of greater knowledge on the phenotypic spectrum of rare chromosomal disorders. This knowledge is extremely valuable for rare diseases, where available information remains limited even though, as also argued before by Schumacher et al ^14^, this information could improve treatment and surveillance and facilitate future research. Collecting data directly from parents recruited via social media results in new opportunities for rare disease research, including on topics that have not been reported before.

## Conclusions

We have shown that phenotype data collected directly from parents of children with a rare disease is highly consistent with data extracted from the medical files on the same individuals. We have also demonstrated that collecting phenotype data via the Chromosome 6 Questionnaire assembles more data on phenotypic characteristics than is presented in the literature. Finally, using social media to recruit participants and collect data directly from rare disease parents is a very reliable and rewarding way to generate more knowledge on rare diseases and unravel the full phenotypic spectrum of a rare disorder.

## Data Availability

All data produced in the present study are available upon reasonable request to the authors

## List of abbreviations

IADL: Lawton Instrumental Activities of Daily Living
DQ: developmental quotient

## Declarations

### Ethics approval and consent to participate

The accredited Medical Ethics Review Committee of the University Medical Center Groningen waived full ethical evaluation because, following Dutch guidelines, no ethical approval is necessary if already available medical information is used anonymously and no extra tests have to be performed.

### Consent for publication

Consent for publication was obtained from all participants.

### Availability of data and materials

Data collected from individuals with terminal 6q deletions in the parent cohort was submitted to the DECIPHER database (decipher.sanger.ac.uk), IDs 489709–489743.

### Authors’ contributions

Conceptualization: AE, BF, CMAR; Data curation: AE, BF; Formal analysis: AE, BF, NFS; Funding acquisition: AE, CMAR; Investigation: AE, BF, CMAR; Methodology: AE, AVR, CMAR; Project administration: AE, BF; Resources: AE, BF, CMAR; Software: AE, MAS; Supervision: CMAR; Visualization: AE; Writing – original draft: AE; Writing – review & editing: AE, BF, ER, NFS, MAS, MP, WSK, AVR, CMAR.

## Acknowledgements

We would like to thank all the children and families for their participation. We also thank Kate Mc Intyre for editing the manuscript, Robert Warmerdam for input on the statistical analysis, the members of the MOLGENIS team at the Groningen Genomic Coordination Center, in particular Mariska Slofstra, for their support on the online Chromosome 6 Questionnaire and the Chromosome 6 Project’s advisory board for their input. Our special thanks go to Pauline Bouman, our contact for the Chromosome 6 Facebook group.

## Additional File 1 .pdf

### Supplementary Tables

Table S1. Country of origin of invited and test panel parents

Table S2. Number of pages collected from medical files

Table S3. Number of health professionals from who medical files were received

Table S4. Number of follow-up measurements for growth available per individual

Table S5. Consistency main questions

Table S6. Consistency sub-questions

Table S7. Number of answers unknown from medical files data

Table S8. Type of inconsistency per question category

Table S9. Type and number of inconsistencies per test panel parent

Table S10. Number of questions for which the availability of data was significantly, or not significantly different when collected from parents compared to literature

Table S11. All p-values main questions

Table S12. All p-values sub-questions

### Supplementary Figures

Figure S1. Flowchart for test panel selection

Figure S2. Flowchart inclusion individuals data availability study

Figure S3. Total answers checked for consistency and distribution of consistent and inconsistent answers per test parent

### Supplementary Information

Supplementary Information S1. List of included questions of Chromosome 6 Questionnaire

