## Supplementary files for "Parent-reported phenotype data on chromosome 6 aberrations collected via an online questionnaire: data consistency and data availability"

### Additional File 1: Supplementary files

**Supplementary Tables**

Table S1. Country of origin of invited and test panel parents
Table S2. Number of pages collected from medical files
Table S3. Number of health professionals from who medical files were received
Table S4. Number of follow-up measurements for growth available per individual
Table S5. Consistency main questions
Table S6. Consistency sub-questions
Table S7. Number of answers unknown from medical files data
Table S8. Type of inconsistency per question category
Table S9. Type and number of inconsistencies per test panel parent
Table S10. Number of questions for which the availability of data was significantly, or not significantly different when collected from parents compared to literature
Table S11. All p-values main questions
Table S12. All p-values sub-questions

**Supplementary Figures**

Figure S1. Flowchart for test panel selection
Figure S2. Flowchart inclusion individuals data availability study
Figure S3. Total answers checked for consistency and distribution of consistent and inconsistent answers per test parent

**Supplementary Information**

Supplementary Information S1. List of included questions of Chromosome 6 Questionnaire

**Table S1.** Country of origin of invited and test panel parents

| Country of origin | Number of parents invited | Number of parents in test panel |
|---|---|---|
| United Kingdom | 7 | 4 |
| Ireland | 1 | 1 |
| United States of America | 28 | 11 |
| Canada | 2 | 1 |
| Australia | 4 | 2 |
| New Zealand | 1 | 1 |

**Table S2.** Number of pages collected from medical files

| Total number of pages received per patient median (range) | Number of health professionals per patient median (range) | Average number of pages per health professional per patient median (range) |
|---|---|---|
| 47.5 (15–121) | 4 (2–11) | 10.4 (3.4–43) |

**Table S3.** Number of health professionals from whom medical files were received

| Health professionals from whom medical files were received | Number of individuals for whom medical files were received |
|---|---|
| Geneticist | 18 |
| Neurologist | 14 |
| Paediatrician | 10 |
| General Practitioner | 9 |
| Ophthalmologist | 9 |
| Otorhinolaryngologist | 5 |
| Speech therapist | 4 |
| Gastroenterologist | 4 |
| Cardiologist | 3 |
| Occupational therapist | 3 |
| Orthopaedics | 3 |
| Physiotherapist | 2 |
| Pulmonologist | 2 |
| Neurosurgeon | 1 |
| Endocrinologist | 1 |
| Psychologist | 1 |
| Paediatric Surgeon | 1 |
| General surgeon | 1 |
| Infectious Disease specialist | 1 |
| Audiologist | 1 |

Information from the General Practitioner often also included letters from other health professionals not mentioned in this table, for example trauma surgeons, plastic surgeons, metabolic specialists and development specialists.

**Table S4.** Number of follow-up measurements for growth available per individual

| Follow-up measurements | Parent data Median (range) | Medical file data Median (range) |
|---|---|---|
| Height | 3.5 (0–5) | 1.5 (1–4) |
| Weight | 4 (0–5) | 2 (1–4) |
| Head circumference | 1.5 (0–5) | 1 (0–3) |

**Table S5.** Consistency main questions

| | Consistency main questions | | | Answer unknown | |
|---|---|---|---|---|---|
| | Total answers | Consistent/observed (+ probably consistent/observed)* | Consistency – maximum consistency* | Parent data | Medical files data |
| **Congenital malformations** | **740** | **130/167 (627/664)** | **78–94%** | **3** | **570** |
| Physical aspects | 580 | 89/115 (492/518) | 77–95% | 3 | 462 |
| *Head* | 100 | 29/29 (91/91) | 100% | 3 | 68 |
| *Face* | 160 | 20/26 (131/137) | 77–96% | 0 | 134 |
| *Trunk* | 180 | 23/31 (166/174) | 74–95% | 0 | 149 |
| *Limbs* | 80 | 14/21 (54/61) | 67%–89% | 0 | 59 |
| *Skin* | 60 | 3/8 (50/55) | 38–91% | 0 | 52 |
| Heart and vessels | 40 | 9/10 (37/38) | 90–97% | 0 | 30 |
| Airways and lungs | 60 | 6/8 (49/51) | 75–96% | 0 | 52 |
| Kidneys and genitals | 40 | 14/17 (34/37) | 82–92% | 0 | 23 |
| Brain | 20 | 12/17 (15/20) | 71–75% | 0 | 3 |
| **Medical problems** | **744** | **146/174 (648/676)** | **84–96%** | **0** | **570** |
| Senses | 100 | 36/48 (79/91) | 75–87% | 0 | 52 |
| *Visual* | 60 | 25/35 (47/57) | 71–82% | 0 | 25 |
| *Hearing* | 40 | 11/13 (32/34) | 85–94% | 0 | 27 |
| Feeding and the gut | 160 | 27/31 (142/146) | 87–97% | 0 | 129 |
| *Feeding* | 20 | 9/9 (13/13) | 100% | 0 | 11 |
| *Gut* | 140 | 18/22 (129/133) | 82–97% | 0 | 118 |
| Heart and vessels | 40 | 1/1 (39/39) | 100% | 0 | 39 |
| Blood, hormones and immune system | 204 | 24/28 (184/188) | 86–98% | 0 | 176 |
| *Blood* | 60 | 4/5 (57/58) | 80–98% | 0 | 55 |
| *Hormones*[&] | 104 | 15/17 (101/103) | 88–98% | 0 | 87 |
| *Immune system* | 40 | 5/6 (26/27) | 83–96% | 0 | 34 |
| Neurology and Epilepsy | 240 | 58/66 (204/212) | 88–96% | 0 | 174 |
| *Neurology* | 220 | 47/55 (186/194) | 85–96% | 0 | 165 |
| *Epilepsy* | 20 | 11/11 (18/18) | 100% | 0 | 9 |
| **Behaviour** | **60** | **8/13 (45/50)** | **62–90%** | **0** | **47** |
| Diagnosis | 40 | 4/6 (32/34) | 67–94% | 0 | 34 |
| Sleeping | 20 | 4/7 (13/16) | 57–81% | 0 | 13 |
| **Growth measurements** | **81** | **50/53 (na)** | **94%** | **1** | **27** |
| Birth measurements | 48 | 19/20 (na) | 95% | 1 | 27 |
| *Birth weight* | 19 | 9/9 (na) | 100% | 1 | 9 |
| *Birth length* | 18 | 8/8 (na) | 100% | 0 | 10 |
| *Head circumference birth* | 11 | 2/3 (na) | 67% | 0 | 8 |
| Follow-up measurements | 33 | 31/33 (na) | 94% | na | na |
| *Height* | na | 12/13 (na) | 92% | na | na |
| *Weight* | na | 12/13 (na) | 92% | na | na |
| *Head circumference* | na | 7/7 (na) | 100% | na | na |
| **Development** | **438** | **109/114 (192/197)** | **96–97%** | **2** | **322** |
| IQ | 40 | 21/23 (27/29) | 91–93% | 0 | 17 |
| Milestones | 398 | 88/91 (165/168) | 97–98% | 2 | 305 |
| *Gross motor* | 160 | 52/52 (61/61) | 100% | 0 | 108 |
| *Fine motor* | 100 | 4/5 (39/40) | 80–98% | 0 | 95 |
| *Communication* | 100 | 30/32 (43/45) | 94–96% | 0 | 68 |
| *Toilet training*[#] | 38 | 2/2 (22/22) | 100% | 2 | 34 |
| **Total** | **2063** | **443/521 (1562/1640)** | **85–95%** | **6** | **1536** |

*Including the probably consistent answers, see Methods.
[&]For 16 individuals, the question about puberty was not applicable due to young age
[#]For 1 individual, the questions about toilet training milestones were not completed.
na = not applicable

**Table S6.** Consistency sub questions

| | Consistency sub questions | | | Answer unknown | |
|---|---|---|---|---|---|
| | Total answers | consistent/observed (+ partly consistent/observed)* | % consistency – maximum consistency* | Parent data | Medical file data |
| **Congenital malformations** | **85** | **67/79 (79/79)** | **85–100%** | **0** | **6** |
| Physical aspects | 51 | 45/47 (47/47) | 96–100% | 0 | 4 |
| *Head* | 7 | 7/7 | 100% | 0 | 0 |
| *Face* | 10 | 9/10 (10/10) | 90–100% | 0 | 0 |
| *Trunk* | 8 | 8/8 | 100% | 0 | 0 |
| *Limbs* | 19 | 16/16 | 100% | 0 | 3 |
| *Skin* | 7 | 5/6 (6/6) | 83–100% | 0 | 1 |
| Heart and vessels | 3 | 2/3 (3/3) | 67–100% | 0 | 0 |
| Airways and lungs | 11 | 10/11 (11/11) | 91–100% | 0 | 0 |
| Kidneys and genitals | 8 | 6/7 (7/7) | 86–100% | 0 | 1 |
| Brain | 12 | 4/11 (11/11) | 36–100% | 0 | 1 |
| **Medical problems** | **115** | **49/78 (75/78)** | **63–96%** | **1** | **36** |
| Senses | 38 | 16/22 (22/22) | 73–100% | 1 | 15 |
| *Visual* | 18 | 11/17 (17/17) | 65–100% | 1 | 0 |
| *Hearing* | 20 | 5/5 | 100% | 0 | 15 |
| Feeding and the gut | 21 | 9/21 (21/21) | 43–100% | 0 | 0 |
| *Feeding* | 12 | 3/12 (12/12) | 25–100% | 0 | 0 |
| *Gut* | 9 | 6/9 (9/9) | 67–100% | 0 | 0 |
| Heart and vessels | 0 | 0/0 | | 0 | 0 |
| Blood, hormones and immune system | 2 | 2/2 | 100% | 0 | 0 |
| *Blood* | 0 | 0/0 | | 0 | 0 |
| *Hormones** | na | na | na | na | na |
| *Immune system* | 2 | 2/2 | 100% | 0 | 0 |
| Neurology and epilepsy | 54 | 22/33 (30/33) | 67–91% | 0 | 21 |
| *Neurology* | na | na | na | na | na |
| *Epilepsy* | 54 | 22/33 (30/33) | 67–91% | 0 | 21 |
| **Behaviour** | **6** | **4/5 (5/5)** | **80–100%** | **1** | **0** |
| Diagnosis | 3 | 3/3 | 100% | 0 | 0 |
| Sleeping | 3 | 1/2 (2/2) | 50–100% | 1 | 0 |
| **Development** | **89** | **36/41** | **88%** | **19** | **29** |
| IQ | 9 | 1/1 | 100% | 4 | 4 |
| Milestone | 80 | 35/40 | 88% | 15 | 25 |
| *Gross motor* | 58 | 30/34 | 88% | 9 | 15 |
| *Fine motor* | 0 | 0/0 | | 0 | 0 |
| *Communication* | 20 | 5/6 | 83% | 4 | 10 |
| *Toilet training* | 2 | 0/0 | | 2 | 0 |
| **Total** | **295** | **156/203 (195/203)** | **77–96%** | 21 | 71 |

na = not applicable, this subcategory did not contain sub-questions.

*including the partly consistent answers, see Methods

**Table S7.** Number of answers unknown from medical files data

| | Number of unknown answers for which parents reported absent | Number of unknown answers for which parents reported present |
|---|---|---|
| Congenital malformations | 497 | 73 |
| Medical problems | 502 | 68 |
| Behaviour | 37 | 10 |
| Development | 83 | 239 |
| **Total** | **1119** | **390** |

**Table S8.** Type of inconsistency per question category

| | Major/total answers | Minor/partly/total answers |
|---|---|---|
| **Congenital malformations** | **37/167** | **0/12/79** |
| Airways and lungs | 2/8 | 0/1/11 |
| Brain | 5/17 | 0/7/11 |
| Heart and vessels | 1/10 | 0/1/3 |
| Kidneys and genitals | 3/17 | 0/1/7 |
| Physical aspects | 26/115 | 0/2/47 |
| *Face* | 6/26 | 0/1/10 |
| *Head* | 0/29 | 0/0/7 |
| *Limbs* | 7/21 | 0/0/16 |
| *Skin* | 5/8 | 0/1/6 |
| *Trunk* | 8/31 | 0/0/8 |
| **Medical problems** | **21/142** | **10/26/110** |
| Blood, hormones and immune system | 4/28 | 0/0/2 |
| *Blood* | 1/5 | 0/0/0 |
| *Hormones* | 2/17 | na |
| *Immune system* | 1/6 | 0/0/2 |
| Epilepsy and neurology | 1/34 | 10/8/65 |
| *Epilepsy* | 0/11 | 3/8/33 |
| *Neurology* | 1/23 | 7/0/32 |
| Feeding and the gut | 4/31 | 0/12/21 |
| *Feeding* | 0/9 | 0/9/12 |
| *Gut* | 4/22 | 0/3/9 |
| Heart and vessels | 0/1 | 0/0/0 |
| Senses | 12/48 | 0/6/22 |
| *Hearing* | 2/13 | 0/0/5 |
| *Visual* | 10/35 | 0/6/17 |
| **Growth measurements** | **3/53** | **na** |
| Birth measurements | 1/20 | na |
| *Birth weight* | 0/9 | na |
| *Birth length* | 0/8 | na |
| *Head circumference birth* | 1/3 | na |
| Follow-up measurements | 2/33 | na |
| *Height* | 1/13 | na |
| *Weight* | 1/13 | na |
| *Head circumference* | 0/7 | na |
| **Behaviour** | **5/13** | **0/1/5** |
| Diagnosis | 2/6 | 0/0/3 |
| Sleeping | 3/7 | 0/1/2 |
| **Development** | **5/114** | **5/na/41** |
| IQ test | 2/23 | 0/na/1 |
| Milestone | 3/91 | 5/na/40 |
| *Gross motor* | 0/52 | 4/na/34 |
| *Fine motor* | 1/5 | 0/na/0 |
| *Communication* | 2/32 | 1/na/6 |
| *Toilet training* | 0/2 | 0/na/0 |
| **Total** | **71/489** | **15/39/235** |

**Table S9.** Type and number of inconsistencies per test panel parent

| | Consistent/ observed | consistency | Total inconsistent | Major | Minor | Partly |
|---|---|---|---|---|---|---|
| **1** | 39/49 | 80% | 10 | 6 | 1 | 3 |
| **2** | 41/51 | 80% | 10 | 8 | 1 | 1 |
| **3** | 28/31 | 90% | 3 | 3 | 0 | 0 |
| **4** | 39/48 | 81% | 9 | 2 | 0 | 7 |
| **5** | 28/37 | 76% | 9 | 4 | 1 | 4 |
| **6** | 14/17 | 82% | 3 | 3 | 0 | 0 |
| **7** | 20/20 | 100% | 0 | 0 | 0 | 0 |
| **8** | 31/35 | 89% | 4 | 1 | 0 | 3 |
| **9** | 30/33 | 91% | 3 | 3 | 0 | 0 |
| **10** | 18/22 | 82% | 4 | 2 | 1 | 1 |
| **11** | 24/26 | 92% | 2 | 2 | 0 | 0 |
| **12** | 36/42 | 86% | 6 | 4 | 2 | 0 |
| **13** | 23/31 | 74% | 8 | 2 | 2 | 4 |
| **14** | 36/41 | 88% | 5 | 2 | 0 | 3 |
| **15** | 36/42 | 86% | 6 | 5 | 0 | 1 |
| **16** | 32/42 | 76% | 10 | 5 | 3 | 2 |
| **17** | 44/59 | 75% | 15 | 8 | 3 | 4 |
| **18** | 27/32 | 84% | 5 | 1 | 0 | 4 |
| **19** | 20/28 | 71% | 8 | 8 | 0 | 0 |
| **20** | 33/38 | 87% | 5 | 2 | 1 | 2 |

This data is visualised in Figure 2 and Figure S3.

**Table S10.** Number of questions for which the availability of data was significantly (or not significantly) different when collected from parents compared to literature

| | Main questions | Sub-questions |
|---|---|---|
| | **Significant more data from parents/ significant more data from literature/ no significant difference (α=0.05)** | **Significant more data from parents/ significant more data from literature/ no significant difference (α=0.05)** |
| **Congenital malformations** | **37/0/1** | **1/1/24** |
| Airways and lungs | 3/0/0 | 0/0/3 |
| Brain | 0/0/1 | 0/1/0 |
| Heart and vessels | 2/0/0 | 0/0/2 |
| Kidneys and genitals | 3/0/0 | 0/0/3 |
| Physical aspects | 29/0/0 | 1/0/16 |
| *Face* | 8/0/0 | 0/0/3 |
| *Head* | 5/0/0 | 0/0/2 |
| *Limbs* | 4/0/0 | 1/0/3 |
| *Skin* | 3/0/0 | 0/0/3 |
| *Trunk* | 9/0/0 | 0/0/5 |
| **Medical problems** | **38/0/0** | **2/0/19** |
| Blood, hormones and immune system | 11/0/0 | 0/0/4 |
| *Blood* | 3/0/0 | 0/0/3 |
| *Hormones* | 6/0/0 | na |
| *Immune system* | 2/0/0 | 0/0/1 |
| Epilepsy and neurology | 12/0/0 | 1/0/3 |
| *Epilepsy* | 1/0/0 | 1/0/3 |
| *Neurology* | 11/0/0 | na |
| Feeding and the gut | 8/0/0 | 1/0/5 |
| *Feeding* | 1/0/0 | 1/0/1 |
| *Gut* | 7/0/0 | 0/0/4 |
| Heart and vessels | 2/0/0 | 0/0/2 |
| Senses | 5/0/0 | 0/0/5 |
| *Hearing* | 2/0/0 | 0/0/2 |
| *Visual* | 3/0/0 | 0/0/3 |
| **Behaviour** | **3/0/0** | **0/0/3** |
| Diagnosis | 2/0/0 | 0/0/2 |
| Sleeping | 1/0/0 | 0/0/1 |
| **Growth measurements** | **6/0/0** | **na** |
| Birth measurements | 3/0/0 | na |
| Follow-up measurements | 3/0/0 | na |
| **Development** | **29/0/1** | **2/0/19** |
| IQ | 0/0/2 | 0/0/1 |
| Milestone | 20/0/0 | 2/0/18 |
| *Gross motor* | 8/0/0 | 0/0/8 |
| *Fine motor* | 5/0/0 | 0/0/5 |
| *Communication* | 5/0/0 | 2/0/3 |
| *Toilet training* | 2/0/0 | 0/0/2 |
| Adult functioning | 8/0/0 | na |
| **Total** | **113/0/2** | **5/1/65** |

na= not applicable

**Table S11.** All p-values main questions

| Main question | Parent known/total | Literature known/total | Chi-square (Fisher's exact test*) (α=0.05) |
|---|---|---|---|
| An abnormal head size | 34/34 | 18/39 | **<0.0001** |
| Abnormality of the fontanelles | 24/34 | 1/39 | **<0.0001** |
| Plagiocephaly (asymmetric skull) | 32/34 | 1/39 | **<0.0001** |
| Craniosynostosis (too early closure of the sutures of the skull) | 30/34 | 0/39 | **<0.0001** |
| Encephalocele (skull defect with protrusion of the brain and/or membranes) | 21/34 | 0/39 | **<0.0001** |
| Hypertelorism (widely spaced eyes) | 32/34 | 19/39 | **<0.0001** |
| Ptosis (drooping) of the upper eyelid | 33/34 | 2/39 | **<0.0001** |
| Blocked or absent tear ducts (nasolacrimal ducts) | 31/34 | 1/39 | **<0.0001** |
| An abnormal lip or palate | 34/34 | 15/39 | **<0.0001** |
| Dental (tooth) abnormalities | 34/34 | 3/39 | **<0.0001** |
| Skin pit or skin tag near the ears | 33/34 | 0/39 | **<0.0001** |
| Abnormality of the outer ear (malformed or dysplastic) | 34/34 | 19/39 | **<0.0001** |
| Abnormality of the auditory canal | 32/34 | 1/39 | **<0.0001** |
| Underdeveloped or absent collarbones (clavicles) | 30/34 | 1/39 | **<0.0001** |
| Abnormality of the ribs | 30/34 | 3/39 | **<0.0001** |
| An abnormal shape of the thorax | 30/34 | 3/39 | **<0.0001** |
| Extra nipples | 30/34 | 0/39 | **<0.0001** |
| An abdominal wall defect | 34/34 | 2/39 | **<0.0001** |
| Abnormal curvature of the back/spine | 32/34 | 5/39 | **<0.0001** |
| Abnormal vertebrae | 30/34 | 3/39 | **<0.0001** |
| Spina bifida | 31/34 | 2/39 | **<0.0001** |
| A sacral dimple (small depression in the skin, located just above the buttocks) | 33/34 | 2/39 | **<0.0001** |
| Hypermobility of the joints | 34/34 | 14/39 | **<0.0001** |
| Contractures of the joints (limited movement of joints due to fixation) | 30/34 | 0/39 | **<0.0001** |
| Particular features of the hands or fingers | 34/34 | 4/39 | **<0.0001** |
| Particular features of the feet or toes | 34/34 | 8/39 | **<0.0001** |
| Abnormalities of the skin | 32/34 | 5/39 | **<0.0001** |
| Abnormal hair | 30/34 | 2/39 | **<0.0001** |

| | | | |
|---|---|---|---|
| Abnormalities of the nails | 34/34 | 1/39 | **<0.0001** |
| Is there a congenital heart defect | 34/34 | 10/39 | **<0.0001** |
| Does your child have cardiomyopathy | 34/34 | 6/39 | **<0.0001** |
| Are there any nasal problems | 33/34 | 0/39 | **<0.0001** |
| Does your child have breathing, airway or lung problems | 34/34 | 1/39 | **<0.0001** |
| Does your child have a diaphragmatic problem | 34/34 | 0/39 | **<0.0001** |
| Does your child have renal or urinary tract problems | 34/34 | 4/39 | **<0.0001** |
| Does your son have abnormalities of the genitals | 24/34 | 5/39 | **<0.0001** |
| Does your daughter have abnormalities of the genitals | 27/34 | 1/39 | **<0.0001** |
| Were there abnormalities of the brain seen on imaging (MRI, CT, ultrasound) | 27/34 | 36/39 | 0.1099 (0.1725) |
| Does your child have any problems with his/her eyes | 33/34 | 12/39 | **<0.0001** |
| Does your child have abnormal (involuntary) eye movements | 34/34 | 19/39 | **<0.0001** |
| Are there special characteristics of the eyes (cataract, coloboma, retinitis pigmentosa, glaucoma, anophthalmia, etc.) | 34/34 | 7/39 | **<0.0001** |
| Are there congenital abnormalities of the middle and/or inner ear | 32/34 | 2/39 | **<0.0001** |
| Does your child have a hearing impairment | 34/34 | 4/39 | **<0.0001** |
| Has your child had any problems in eating, drinking or swallowing | 34/34 | 5/39 | **<0.0001** |
| Oesophageal atresia (congenitally interrupted oesophagus) | 34/34 | 1/39 | **<0.0001** |
| A tracheo-oesophageal fistula (abnormal connection (fistula) between the oesophagus and the trachea) | 34/34 | 1/39 | **<0.0001** |
| Gastro-oesophageal or acid reflux (GERD) | 34/34 | 0/39 | **<0.0001** |
| An abnormality of the liver or poor liver function | 34/34 | 0/39 | **<0.0001** |
| An abnormality of the gallbladder or bile ducts | 34/34 | 0/39 | **<0.0001** |
| Bowel or defecation problems | 34/34 | 1/39 | **<0.0001** |
| Anal problems | 32/34 | 1/39 | **<0.0001** |
| Does your child have arrhythmia | 31/34 | 4/39 | **<0.0001** |
| Does your child have abnormal blood pressure | 30/34 | 0/39 | **<0.0001** |
| Are there abnormalities of the number of blood cells | 30/34 | 0/39 | **<0.0001** |
| Are there abnormalities in blood clotting | 30/34 | 0/39 | **<0.0001** |
| Is there an abnormality of the spleen | 30/34 | 0/39 | **<0.0001** |
| Growth hormone (too little/too much/no problem) | 34/34 | 1/39 | **<0.0001** |
| Thyroid hormone (too little/too much/no problem) | 34/34 | 0/39 | **<0.0001** |
| Adrenal hormones (too little/too much/no problem) | 34/34 | 0/39 | **<0.0001** |
| Diabetes mellitus | 34/34 | 0/39 | **<0.0001** |

| | | | |
|---|---|---|---|
| Diabetes insipidus (condition characterised by excessive thirst and excretion of large amounts of severely dilute urine) | 30/34 | 0/39 | **<0.0001** |
| Did your child enter puberty spontaneously | 7/34 | 1/39 | **0.0139 (0.0215)** |
| Does or did your child have recurrent infections | 30/34 | 1/39 | **<0.0001** |
| Does your child have a problem with his/her immune system | 30/34 | 0/39 | **<0.0001** |
| Facial palsy | 30/34 | 0/39 | **<0.0001** |
| Hypotonia or low muscle tone | 32/34 | 28/39 | **0.0129** |
| Hypertonia or excessive muscle tone | 27/34 | 2/39 | **<0.0001** |
| Spasticity (combination of paralysis, increased tendon reflex activity, hypertonia, excessive muscle contraction) | 30/34 | 0/39 | **<0.0001** |
| Hyperreflexia (overactive or over-responsive tendon reflexes) | 30/34 | 2/39 | **<0.0001** |
| Areflexia/hyporeflexia (below normal or absent tendon reflexes ) | 30/34 | 2/39 | **<0.0001** |
| Myopathy (muscular disease resulting in muscular weakness) | 30/34 | 1/39 | **<0.0001** |
| Ataxia (poor coordination of muscle movements) | 30/34 | 2/39 | **<0.0001** |
| Torticollis (twisted neck due to hypertonia of neck muscles) | 31/34 | 0/39 | **<0.0001** |
| Is there an abnormality of the spinal cord (tethered cord) | 34/34 | 8/39 | **<0.0001** |
| Does your child have migraine headaches | 30/34 | 0/39 | **<0.0001** |
| Has your child ever had convulsions, fits or seizures | 34/34 | 31/39 | **0.0051 (0.0059)** |
| Was there a behavioural diagnosis made by a (child) psychiatrist or psychologist? | 30/34 | 3/39 | **0.0001** |
| Does your child have a mood disorder | 30/34 | 0/39 | **<0.0001** |
| Does your child have problems with sleeping | 33/34 | 2/39 | **<0.0001** |
| Weight at birth | 33/34 | 11/39 | **<0.0001** |
| Length at birth | 27/34 | 10/39 | **<0.0001** |
| Head circumference at birth | 18/34 | 10/39 | **0.0167 (0.0291)** |
| Height follow-up | 31/34 | 12/39 | **<0.0001** |
| Weight follow-up | 33/34 | 10/39 | **<0.0001** |
| Head circumference follow-up | 29/34 | 22/39 | **0.0073 (0.0102)** |
| Is there developmental delay | 25/25 | 31/31 | 1 (1) |
| Has an IQ, DQ or developmental test been done | 34/34 | 6/39 | **<0.0001** |
| Reach for objects | 33/34 | 0/39 | **<0.0001** |
| Roll over from lying on back to lying on tummy | 34/34 | 1/39 | **<0.0001** |
| Sit up unsupported | 34/34 | 3/39 | **<0.0001** |
| Get to a sitting position without help | 34/34 | 5/39 | **<0.0001** |
| Crawl or bottom shuffle | 34/34 | 1/39 | **<0.0001** |
| Pull him- or herself up a standing position | 34/34 | 1/39 | **<0.0001** |

| | | | |
|---|---|---|---|
| Walk holding on to furniture (e.g. to a table) | 34/34 | 1/39 | **<0.0001** |
| Walk unsupported | 34/34 | 14/39 | **<0.0001** |
| Pick up something, like an object the size of a currant, between thumb and index finger | 33/34 | 0/39 | **<0.0001** |
| Hold a pencil between thumb and index finger | 32/34 | 0/39 | **<0.0001** |
| Colour within the lines | 32/34 | 0/39 | **<0.0001** |
| Build a tower of 6 blocks | 34/34 | 0/39 | **<0.0001** |
| String beads | 34/34 | 0/39 | **<0.0001** |
| Smile | 34/34 | 0/39 | **<0.0001** |
| Make sounds; cooing, gurgle | 33/34 | 1/39 | **<0.0001** |
| Make sounds; babble, copy sounds he or she hears | 33/34 | 2/39 | **<0.0001** |
| Speak his or her first words | 34/34 | 7/39 | **<0.0001** |
| Speak 2-word sentences | 34/34 | 7/39 | **<0.0001** |
| Is your child fully toilet trained (urine and defecation) during the day | 34/34 | 2/39 | **<0.0001** |
| Is your child fully toilet trained (urine and defecation) during the night | 34/34 | 1/39 | **<0.0001** |
| Ability to use telephone[#] | 2/2 | 0/8 | **0.0222** |
| Shopping[#] | 2/2 | 0/8 | **0.0222** |
| Food preparation[#] | 2/2 | 0/8 | **0.0222** |
| Housekeeping[#] | 2/2 | 0/8 | **0.0222** |
| Laundry[#] | 2/2 | 0/8 | **0.0222** |
| Mode of transportation[#] | 2/2 | 0/8 | **0.0222** |
| Responsibility for own medications[#] | 2/2 | 0/8 | **0.0222** |
| Ability to handle finances[#] | 2/2 | 0/8 | **0.0222** |

[#] Questions only applicable to adults (>18 years)

* A Fisher's exact test was performed when applicable, 20% of the expected counts <5

Bold text indicates significant outcomes

**Table S12.** All p-values sub questions

| Main question followed by sub question | Parent known/main question present | Literature known/main question present | Fisher's exact test (α=0.05) |
|---|---|---|---|
| An abnormal head size → What kind (microcephaly/macrocephaly) | 15/15 | 16/16 | 1 |
| Craniosynostosis (too early closure of sutures of the skull) → Which suture(s) | 1/1 | 0/0 | 1 |
| An abnormal lip or palate → Please specify | 2/7 | 0/13 | 0.1105 |
| Dental (tooth) abnormalities → Please specify | 7/10 | 1/3 | 0.5105 |
| Abnormality of the auditory canal → Please specify | 2/3 | 0/0 | 1 |
| Abnormality of the ribs → Please specify | 0/0 | 1/2 | 1 |
| An abnormal shape of the thorax → Please specify | 0/0 | 1/2 | 1 |
| An abdominal wall defect → Please specify | 4/4 | 2/2 | 1 |
| Abnormal curvature of the back/spine → Please specify | 3/3 | 3/3 | 1 |
| Abnormal vertebrae → Please specify | 3/3 | 0/1 | 0.25 |
| Hypermobility of the joints → Please specify | 15/16 | 2/14 | **<0.0001** |
| Contractures of the joints (limited movement of joints due to fixation) → Please specify | 2/4 | 0/0 | 1 |
| Particular features of the hands or fingers → Please specify | 2/7 | 1/3 | 1 |
| Particular features of the feet or toes → Please specify | 5/6 | 7/7 | 0.4615 |
| Abnormalities of the skin → Please specify | 6/8 | 0/4 | 0.0606 |
| Abnormal hair → Please specify | 0/0 | 1/1 | 1 |
| Abnormalities of the nails → Please specify | 0/0 | 0/1 | 1 |
| Is there a congenital heart defect → Please specify | 8/8 | 3/4 | 0.3333 |
| Does your child have cardiomyopathy → Please specify | 1/1 | 0/0 | 1 |
| Are there any nasal problems → Please specify | 1/2 | 0/0 | 1 |
| Does your child have breathing, airway or lung problems → Please specify | 5/7 | 0/1 | 0.375 |
| Does your child have a diaphragmatic problem → Please specify | 2/2 | 0/0 | 1 |
| Does your child have renal or urinary tract problems → Please specify | 4/5 | 1/1 | 1 |
| Does your son have abnormalities of the genitals → Please specify | 0/2 | 1/2 | 1 |
| Does your daughter have abnormalities of the genitals → Please specify | 3/5 | 0/1 | 1 |
| Were there abnormalities of the brain seen on imaging (MRI, CT, ultrasound) → Please specify | 16/22 | 35/35 | **0.0021** |
| Does your child have any problems with his/her eyes → Please specify | 20/24 | 5/6 | 1 |
| Does your child have abnormal (involuntary) eye movements → Please specify | 14/16 | 13/14 | 1 |

| | | | |
|---|---|---|---|
| Are there special characteristics of the eyes (cataract, coloboma, retinitis pigmentosa, glaucoma, anophthalmia, etc.) → Please specify | 4/9 | 1/2 | 1 |
| Are there congenital abnormalities of the middle and/or inner ear → Please specify | 0/4 | 0/0 | 1 |
| Does your child have a hearing impairment → Is there progressive hearing loss | 0/0 | 0/0 | no data |
| Has your child had any problems in eating, drinking or swallowing→ Please specify | 23/25 | 0/3 | **0.0031** |
| Has your child had any problems in eating, drinking or swallowing → Was one or both of the following needed for feeding problems (tube feeding) | 7/25 | 2/3 | 0.2344 |
| An abnormality of the liver or poor liver function → Please specify | 0/1 | 0/0 | 1 |
| An abnormality of the gallbladder or bile ducts → Please specify | 0/0 | 0/0 | no data |
| Bowel or defecation problems → Please specify | 14/14 | 1/1 | 1 |
| Anal problems → Please specify | 3/4 | 1/1 | 1 |
| Does your child have arrhythmia → Please specify | 0/0 | 0/0 | no data |
| Does your child have abnormal blood pressure → Please specify | 1/2 | 0/0 | 1 |
| Are there abnormalities of the number of blood cells → Please specify | 1/1 | 0/0 | 1 |
| Are there abnormalities in blood clotting → Please specify | 1/1 | 0/0 | 1 |
| Is there an abnormality of the spleen → Please specify | 0/0 | 0/0 | no data |
| Does your child have a problem with his/her immune system → Please specify | 0/2 | 0/0 | 1 |
| Has your child ever had convulsions, fits or seizures → What kind of seizures | 17/19 | 17/24 | 0.2575 |
| Has your child ever had convulsions, fits or seizures → Was epilepsy formally diagnosed | 13/19 | 15/24 | 0.7554 |
| Has your child ever had convulsions, fits or seizures → Is or was there medication for epilepsy used | 17/19 | 9/24 | **0.0006** |
| Has your child ever had convulsions, fits or seizures → Has your child ever had a status epilepticus (an epileptic seizure of more than 30 minutes) | 3/19 | 1/24 | 0.306 |
| Was there a behavioural diagnosis made by a (child) psychiatrist or psychologist? → Please specify | 6/7 | 3/3 | 1 |
| Does your child have a mood disorder → Please specify | 2/2 | 0/0 | 1 |
| Does your child have problems with sleeping → What kind of sleeping problems | 13/15 | 1/2 | 0.3309 |
| Has an IQ, DQ or developmental test been done → What was the result of the testing | 10/10 | 6/6 | 1 |
| Reach for objects → Since what age (months) | 28/32 | 0/0 | 1 |
| Roll over from lying on back to lying on tummy → Since what age (months) | 29/32 | 1/1 | 1 |
| Sit up unsupported → Since what age (months) | 30/31 | 2/2 | 1 |
| Sit up unsupported → Since what age (months) | 25/30 | 4/4 | 1 |
| Crawl or bottom shuffle → Since what age (months) | 27/29 | 1/1 | 1 |
| Pull him- or herself up a standing position → Since what age (months) | 24/25 | 0/0 | 1 |
| Walk holding on to furniture (e.g. to a table) → Since what age (months) | 22/23 | 0/0 | 1 |
| Walk unsupported → Since what age (months) | 18/19 | 10/12 | 0.5435 |

| | | | |
|---|---|---|---|
| Pick up something, like an object the size of a currant, between thumb and index finger → Since what age (months) | 21/27 | 0/0 | 1 |
| Hold a pencil between thumb and index finger → Since what age (months) | 11/15 | 0/0 | 1 |
| Colour within the lines → Since what age (months) | 4/15 | 0/0 | 1 |
| Build a tower of 6 blocks → Since what age (months) | 13/15 | 0/0 | 1 |
| String beads → Since what age (months) | 14/16 | 0/0 | 1 |
| Smile → Since what age (weeks) | 31/34 | 0/0 | 1 |
| Make sounds; cooing, gurgle → Since what age (weeks) | 24/31 | 0/1 | 1 |
| Make sounds; babble, copy sounds he or she hears → Since what age (months) | 25/30 | 1/1 | 1 |
| Speak his or her first words → Since what age (months) | 23/25 | 2/5 | **0.0219** |
| Speak 2-word sentences → Since what age (months) | 18/20 | 1/5 | **0.0055** |
| Is your child fully toilet trained (urine and defecation) during the day → Since what age (years) | 10/10 | 0/0 | 1 |
| Is your child fully toilet trained (urine and defecation) during the night → Since what age (years) | 7/7 | 0/0 | 1 |

Bold text indicates significant outcomes. Sub-questions were only shown in the questionnaire when applicable (abnormality present or milestone achieved).

**Figure S1.** Flowchart for test panel selection

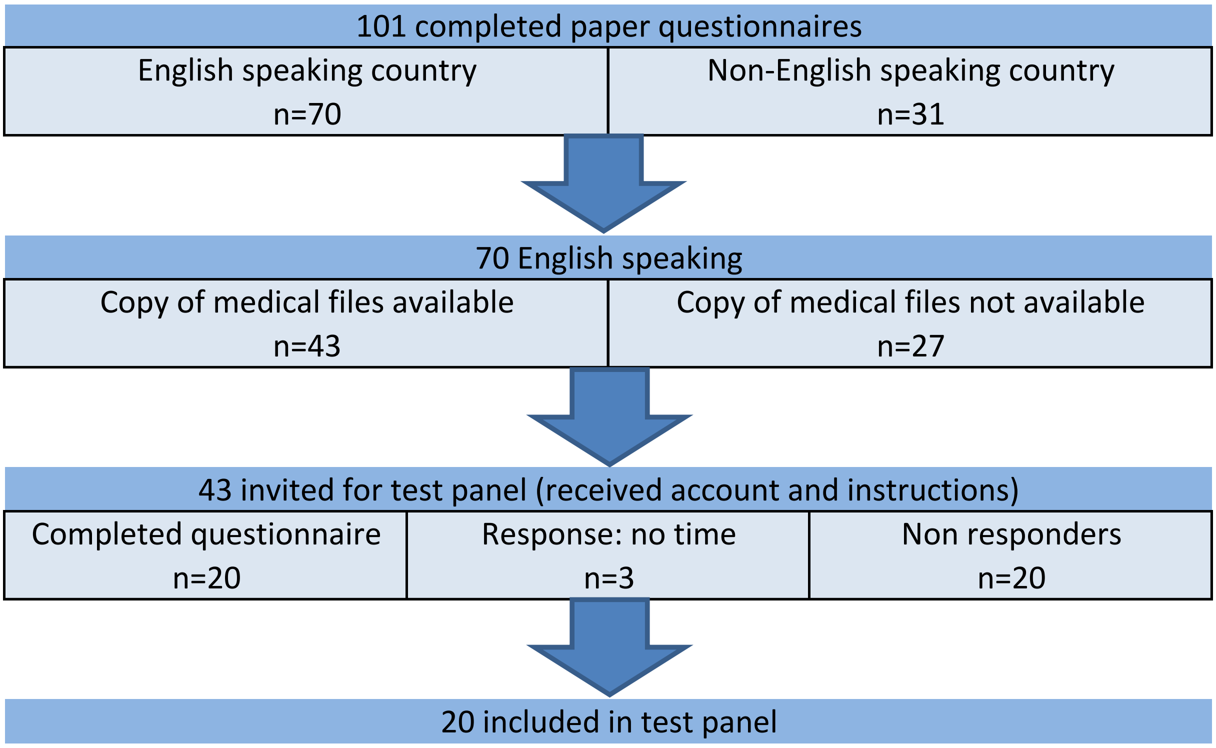


For the parent test panel, 43 parents were invited via email. These parents were selected from the pilot study based on the availability of copies of medical files and on being a native-English speaker. Three parents informed us that they could not be part of the test panel due to time constraints. Twenty parents completed the questionnaire for their child and could be included in the data consistency study.

**Figure S2.** Flowchart inclusion of individuals for data availability study

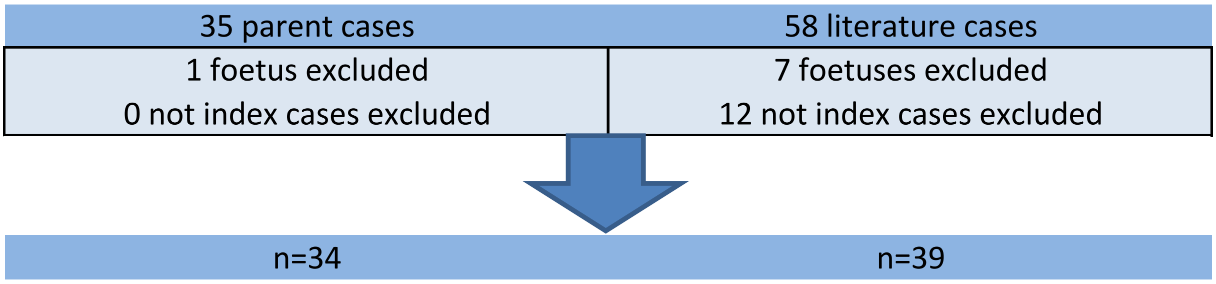

**Figure S3.** Total answers checked for consistency and distribution of consistent and inconsistent answers per test parent

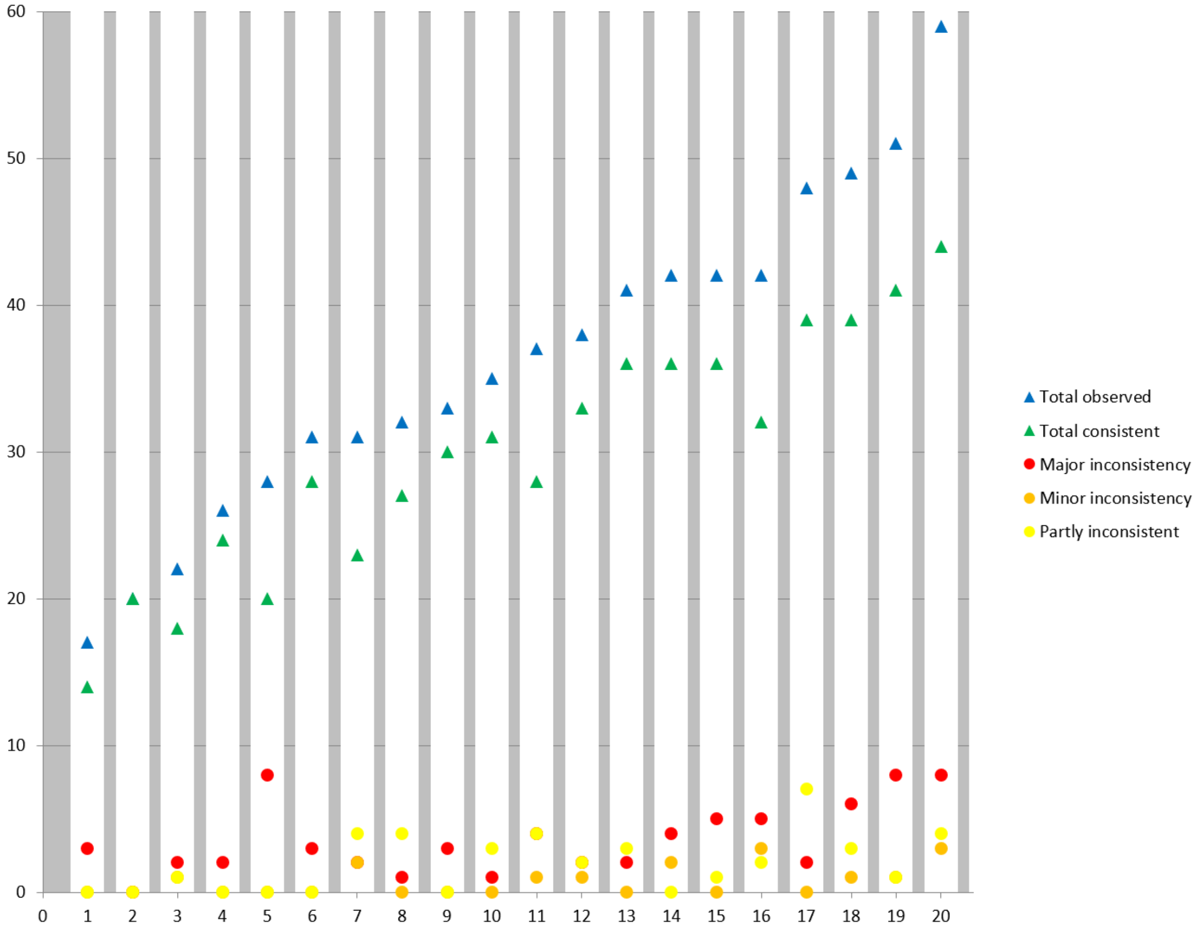


X-axis is the 20 test panel parents. Y-axis is the number of answers.

**Supplementary Information S1.** List of included questions of Chromosome 6 Questionnaire

All included main and sub questions. Sub questions are underlined. Categories and subcategories of questions are in bold.
* = Main question, but considered minor when answers were inconsistent

**Congenital malformations**

**Physical aspects**

***Head***

An abnormal head size

What kind (microcephaly/macrocephaly)

Abnormality of the fontanelles

Plagiocephaly (asymmetric skull)

Craniosynostosis (too early closure of sutures of the skull)

Which suture(s)

Encephalocele (skull defect with protrusion of the brain and/or membranes)

***Face***

Hypertelorism (widely spaced eyes)

Ptosis (drooping) of the upper eyelid

Blocked or absent tear ducts (nasolacrimal ducts)

An abnormal lip or palate

Please specify

Dental (tooth) abnormalities

Please specify

Skin pit or skin tag near the ears

Abnormality of the outer ear (malformed or dysplastic)

Abnormality of the auditory canal

Please specify

***Trunk***

Underdeveloped or absent collarbones (clavicles)

Abnormality of the ribs

Please specify

An abnormal shape of the thorax

Please specify

Extra nipples

An abdominal wall defect

Please specify

Abnormal curvature of the back/spine

Please specify

Abnormal vertebrae

Please specify

Spina bifida

A sacral dimple (small depression in the skin, located just above the buttocks)

***Limbs***

Hypermobility of the joints

Please specify

Contractures of the joints (limited movement of joints due to fixation)

Please specify

Particular features of the hands or fingers

Please specify

Particular features of the feet or toes

Please specify

***Skin***

Abnormalities of the skin

Please specify

Abnormal hair

Please specify

Abnormalities of the nails

Please specify

**Heart and vessels**

Is there a congenital heart defect

Please specify

Does your child have cardiomyopathy

Please specify

**Airways and lungs**

Are there any nasal problems

Please specify

Does your child have breathing, airway or lung problems

Please specify

Does your child have a diaphragmatic problem

Please specify

**Kidneys and genitals**

Does your child have renal or urinary tract problems

Please specify

Does your son have abnormalities of the genitals

Please specify

Does your daughter have abnormalities of the genitals

Please specify

**Brain**

Were there abnormalities of the brain seen on imaging (MRI, CT, ultrasound)

Please specify

**Medical problems**

**Senses**

***Visual***

Does your child have any problems with his/her eyes

Please specify

Does your child have abnormal (involuntary) eye movements

Please specify

Are there special characteristics of the eyes (cataract, coloboma, retinitis pigmentosa, glaucoma, anophthalmia, etc.)

Please specify

***Hearing***

Are there congenital abnormalities of the middle and/or inner ear

Please specify

Does your child have a hearing impairment

Is there progressive hearing loss

**Feeding and the gut**

***Feeding***

Has your child had any problems in eating, drinking or swallowing

Please specify

Was one or both of the following needed for feeding problems (tube feeding)

***Gut***

*Oesophageal atresia (congenitally interrupted oesophagus)

*A tracheo-oesophageal fistula (abnormal connection (fistula) between the oesophagus and the trachea)

Gastro-oesophageal or acid reflux (GERD)

An abnormality of the liver or poor liver function

Please specify

An abnormality of the gallbladder or bile ducts

Please specify

Bowel or defecation problems

Please specify

Anal problems

Please specify

**Heart and vessels**

Does your child have arrhythmia

Please specify

Does your child have abnormal blood pressure

Please specify

**Blood, hormones and immune system**

***Blood***

Are there abnormalities of the number of blood cells

<u>Please specify</u>

Are there abnormalities in blood clotting

<u>Please specify</u>

Is there an abnormality of the spleen

<u>Please specify</u>

***Hormones***

Growth hormone (too little/too much/no problem)

Thyroid hormone (too little/too much/no problem)

Adrenal hormones (too little/too much/no problem)

Diabetes mellitus

Diabetes insipidus (condition characterised by excessive thirst and excretion of large amounts of severely dilute urine)

Did your child enter puberty spontaneously

***Immune system***

Does or did your child have recurrent infections

Does your child have a problem with his/her immune system

<u>Please specify</u>

**Neurology and epilepsy**

***Neurology***

Facial palsy

*Hypotonia or low muscle tone

*Hypertonia or excessive muscle tone

*Spasticity (combination of paralysis, increased tendon reflex activity, hypertonia, excessive muscle contraction)

*Hyperreflexia (overactive or over-responsive tendon reflexes)

*Areflexia/hyporeflexia (below normal or absent tendon reflexes )

Myopathy (muscular disease resulting in muscular weakness)

Ataxia (poor coordination of muscle movements)

Torticollis (twisted neck due to hypertonia of neck muscles)

Is there an abnormality of the spinal cord (tethered cord)

Does your child have migraine headaches

***Epilepsy***

Has your child ever had convulsions, fits or seizures

<u>What kind of seizures</u>

<u>Was epilepsy formally diagnosed</u>

<u>Is or was there medication for epilepsy used</u>

<u>Has your child ever had a status epilepticus (an epileptic seizure of more than 30 minutes)</u>

**Behaviour**

**Behavioural diagnosis**

Was there a behavioural diagnosis made by a (child) psychiatrist or psychologist?

Please specify

Does your child have a mood disorder

Please specify

**Sleeping problems**

Does your child have problems with sleeping

What kind of sleeping problems

**Growth measurements**

**Birth measurements**

Weight at birth (grams, ounces)

Length at birth (cm, inches)

Head circumference at birth (xx.x cm, inches)

**Current measurements**

Height (cm, feet, inches)

Weight (kg, pounds, ounces)

Head circumference (xx.x cm, inches)

**Development**

**IQ**

Is there developmental delay

Has an IQ, DQ or developmental test been done

What was the result of the testing

**Milestones**[a]

***Gross motor milestones***

Reach for objects

Since what age (months)

Roll over from lying on back to lying on tummy

Since what age (months)

Sit up unsupported

Since what age (months)

Get to a sitting position without help

Since what age (months)

Crawl or bottom shuffle

Since what age (months)

Pull him- or herself up a standing position

Since what age (months)

Walk holding on to furniture (e.g. to a table)

Since what age (months)

Walk unsupported

Since what age (months)

***Fine motor milestones***

Pick up something, like an object the size of a currant, between thumb and index finger

Since what age (months)

Hold a pencil between thumb and index finger

Since what age (months)

Colour within the lines

Since what age (months)

Build a tower of 6 blocks

Since what age (months)

String beads

Since what age (months)

***Communication milestones***

Smile

Since what age (weeks)

Make sounds: cooing, gurgle

Since what age (weeks)

Make sounds: babble, copy sounds he or she hears

Since what age (months)

Speak his or her first words

Since what age (months)

Speak 2-word sentences

Since what age (months)

***Toilet training milestones***

Is your child fully toilet trained (urine and defecation) during the day

Since what age (years)

Is your child fully toilet trained (urine and defecation) during the night

Since what age (years)

**Adult functioning**[b]

Ability to use telephone

Shopping

Food preparation

Housekeeping

Laundry

Mode of transportation

Responsibility for own medications

Ability to handle finances

[a] Developmental milestones were derived from the 'Van Wiechenschema', a developmental scale for children used by most Dutch health professionals[1].
[b] Questions on adult functioning were derived from the Lawton Instrumental Activities of Daily Living (IADL) Scale[2].
